# Identifying Psychiatric Manifestations in Outpatients with Depression and Anxiety: A Large Language Model-Based Approach

**DOI:** 10.1101/2025.01.03.24318117

**Authors:** Shihao Xu, Yiming Yan, Yanli Ding, Feng Li, Shu Zhang, Haoyun Tang, Chao Luo, Yan Li, Hao Liu, Yu Mei, Wenjie Gu, Hong Qiu, Yong Wang, Jianyin Qiu, Tao Yang, Zike Wang, Qing Zhang, Haiyang Geng, Yunyun Han, Jun Shao, Nils Opel, Lidong Bing, Min Zhao, Yifeng Xu, Xun Jiang, Jianhua Chen

## Abstract

**Purpose:** Accurate psychiatric diagnosis and assessment are crucial for effective treatment. However, while current data-driven approaches emphasize diagnostic outcomes, the process of decoding the underlying symptom expressions in patients’ language and mapping them to well-defined psychiatric terminology has received relatively little attention. This study investigates the potential of Large Language Models (LLMs) to automate the identification of diagnostic categories and symptoms from psychiatrist-patient dialogues, to provide interpretable insights and support automatic diagnosis.

**Methods:** We analyzed audio recordings from 1160 psychiatric diagnostic interviews, primarily involving patients with depressive disorder and anxiety disorder. A clinical entities corpus was formed by leveraging clinical annotations in EMRs (e.g., chief complaints, mental status, elements in assessment scales) and widely used assessment scales. LLMs were utilized to identify clinical symptoms, rate assessment scales, and an ensemble learning pipeline was designed to classify diagnostic results and symptoms with 10-fold cross-validation.

**Results:** The system achieved 86.9% accuracy for identifying the appearance of clinical annotations and 74.7% (77.2%) accuracy for identifying anxiety (depression) symptoms. Patients with depression and anxiety, diagnosed using ICD-10 codes, were differentiated with an accuracy of 75.5%. Analysis of LLM-generated features shows that depression cases exhibited prominent markers of anhedonia and decreased volition, whereas anxiety disorders were characterized by tension and an inability to relax.

**Conclusion:** This study demonstrates the potential of integrating LLM technology with linguistic and acoustic features to enhance psychiatric diagnostics. The developed pipeline effectively predicts psychiatric diagnoses and provides interpretable insights, showcasing a valuable tool for clinicians in mental health assessment.

## 1 Introduction

Depression and anxiety disorders represent two of the most prevalent mental health conditions globally. Globally, it is estimated that over 300 million people suffer from major depressive disorders, which is equivalent to 4.4% of the world’s population. A similar number of people suffer from anxiety disorders, often with co-occurring depression [7]. The emerging field of digital phenotyping, which involves the nuanced quantification of human phenotypic expression at the individual level through digital device data, offers a quantitative approach to longitudinal observation [2].

The emerging field of digital phenotyping, characterized by continuous and nuanced quantification of human phenotypic expression at the individual level by leveraging digital device data, provides a quantitative approach for longitudinal observation [2]. Researchers have demonstrated that social signals (e.g., linguistics, speech, etc.) play a crucial role in the diagnosis and assessment of patients with depression and anxiety [37, 21]. In particular, the content of a patient’s speech provides rich information about their mental state, cognitive patterns, and emotional experiences [29, 46]. The linguistic features, topic choices, and narrative structures employed by individuals can offer valuable insights into their psychological well-being [46].

Recent advances in NLP, particularly in LLMs such as GPT [32], Gemini [39], and Qwen [40], demonstrate diverse capabilities in clinical reasoning, social media analysis, and psychiatric education [31], which could potentially provide objective, data-driven insights in psychiatry. Moreover, LLMs are able to process, generate, and respond to natural language inputs, which fit naturally into the NIMH’s Research Domain Criteria (RDoC) framework, which suggests new ways of classifying mental disorders based on dimensions of observable behaviors [30]. In recent psychiatric studies, these LLMs excel at understanding and generating complex linguistic patterns with human-like performance, making them widely explored for social media content analysis [22, 42], treatment performance enhancement [41, 1, 6], chat counselor [27, 25], and supporting clinical decision-making [44, 11] from an evidence-based practice perspective.

Although LLMs demonstrate linguistic understanding and generation, they remain relatively scarce in producing objective digital biomarkers in psychiatry [4]. Studies have shown that the speech of patients with depression and anxiety contains distinctive quantitative verbal and nonverbal digital markers compared to healthy controls [46, 21], but these characteristics often remain too subtle for humans to perceive actionable insights, making their practical application and improvement challenging [13]. LLM is able to generate diagnostic results and provide reasoning steps, benefiting from a large amount of pre-training data. However, the interpretation and alignment of answers or decisions generated by LLM remain challenging [23]. Moreover, most studies on depression and anxiety rely primarily on two data sources: social media and structured clinical reports, and are often constrained by limited data availability [37]. Distinguishing between depression and anxiety in clinical settings remains challenging due to the overlap of symptoms and the high comorbidity rate, with limited research on the discovery of objective biomarkers for both conditions [4]. In addition, during clinical interviews, psychiatrists translate patients’ informal symptom descriptions into professional diagnostic terminology; however, there remains a lack of approaches to automatically and effectively bridge this “semantic gap” between patients and clinicians.

To address these gaps in existing research, we collected a comprehensive dataset of psychiatric interviews at the Shanghai Mental Health Center (SMHC) in China, comprising over 15,000 minutes of speech recordings from 1,160 individual outpatients with 25 different diagnoses. These recordings, primarily featuring patients diagnosed with depression and anxiety disorders, were collected in unstructured real-world environments to ensure ecological validity. To mimic the characteristics of clinical diagnosis, we designed a corpus of clinical indicators that incorporates diagnostic criteria, main complaints, mental status evaluations, and components from assessment scales using the Electronic Medical Records (EMRs) in the SMHC and widely-used assessment scales. Subsequently, we employed the pre-trained LLM to indicate the appearance of a corpus of clinical-related symptoms, rate the components of several assessment scales, and further fine-tuned the LLM with clinical annotations from professional psychiatrists to enhance its understanding of clinical-related concepts. In parallel, we extracted linguistic usage patterns and acoustic features to broaden the spectrum of biomarkers. Through the fusion of these modalities, we constructed an ensemble machine-learning pipeline capable of predicting both outpatient diagnostic groups and symptoms with moderately high accuracies. Moreover, we conducted an in-depth analysis of salient patterns between different diagnostic groups to enhance clinical interpretability. Our results demonstrate that objective cues extracted by the LLM, combined with other behavioral markers, can serve as valuable features for differentiating diagnostic groups and identifying symptom disclosure, potentially enhancing both the efficiency and effectiveness of psychiatric diagnosis and assessment in clinical practice.

## 2 Method

This study collected the audio recording of 1,160 participants between August 2023 and January 2024, in collaboration with the SMHC. The overall pipeline is shown in Figure 1. Firstly, we preprocessed and anonymized the recordings before transcribing them automatically into text and performing manual corrections. Secondly, we collaborated with professional psychiatrists to design a set of clinical entities and leveraged the LLM to identify these concepts using the transcripts as input, enhancing the LLM based on the psychiatrists’ annotations through supervised fine-tuning (SFT). Linguistic and acoustic features were extracted from both the transcripts and the speech. Finally, we utilized different modalities to train an ensemble machine learning pipeline to differentiate diagnostic groups and the major symptoms.

**Fig. 1:**
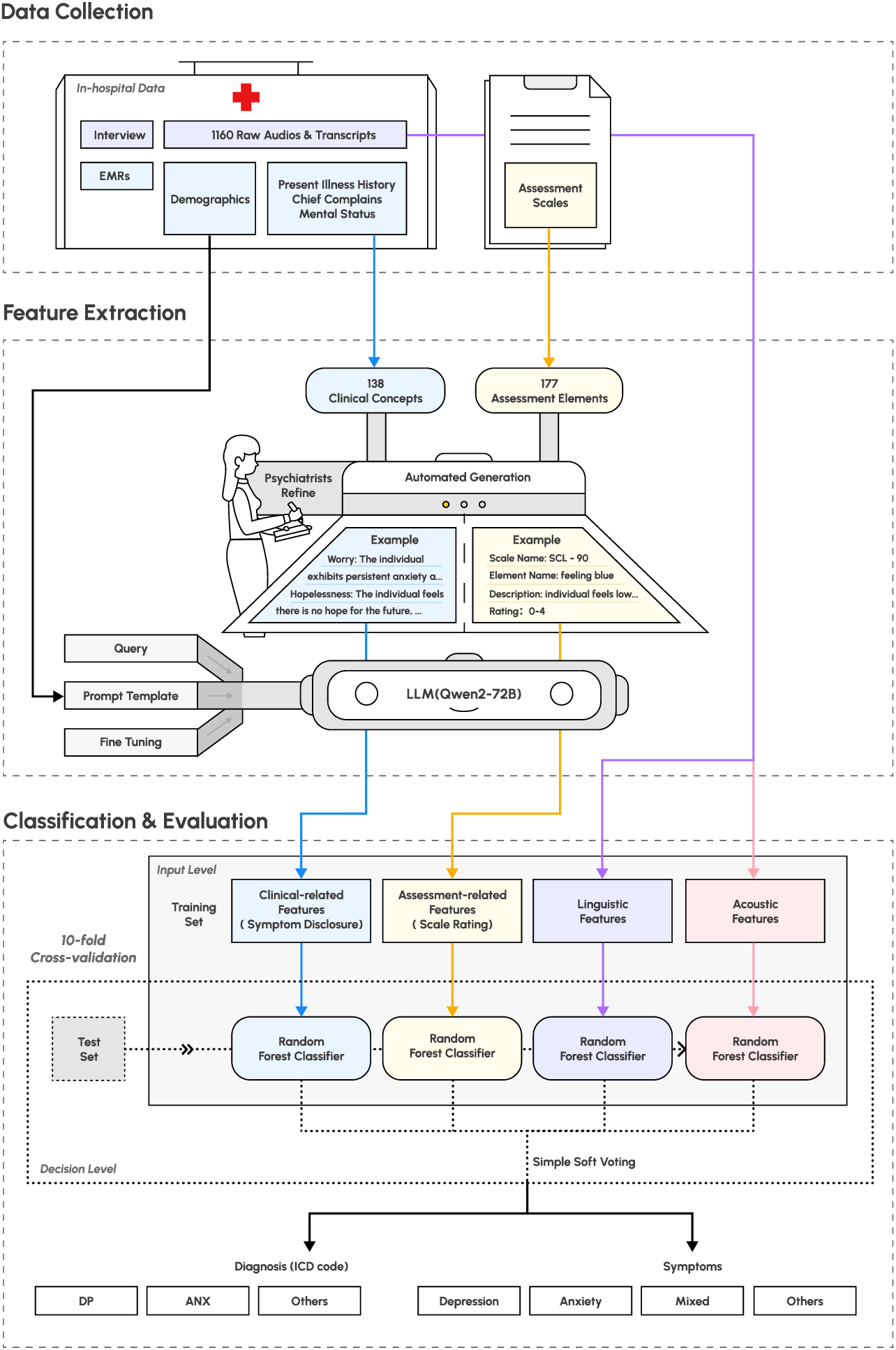
Diagram of the analysis pipeline. The audio recordings were collected during the diagnosis interview for outpatients. We extracted four types of feature sets from the recordings, two of which utilized LLM. These feature sets were utilized to classify different groups of participants and predict the appearance of depression and anxiety symptoms.

### 2.1 Participants

The study sample comprised outpatients from the SMHC who attended daily clinical diagnostic interviews. Participants were aged 12 to 80 years and were fluent in Mandarin. Inclusion criteria required individuals to be capable of providing informed consent and to be free from physical illnesses that could affect their participation. All diagnoses were established using the Chinese version of WHO International Classification of Diseases, Tenth Revision (ICD-10) [20]. The study protocol was approved by the Ethics Committee of the SMHC institutional review board (IRB) to ensure compliance with ethical research standards. Specifically, the recording setup consisted of a microphone placed between the psychiatrist and the participant, connected to a computer. At the beginning of each interview, participants were asked to read a standardized 30-second text passage, followed by the standard diagnostic procedure. All clinical information was documented in the EMR system by the psychiatrists. To protect the privacy of participants, all audio recordings and associated meta-information underwent a thorough manual de-identification process.

### 2.2 Feature Extraction

We extracted a comprehensive clinical entity set to cover the intermediate features that assist psychiatrists in the diagnosis and assessment process: clinical observations and standardized assessment scales, which we designate as clinical-related and assessment-related feature sets. As compensation, we measured the linguistic usage and acoustic characteristics and form as individual feature sets. In the following paragraphs, we will introduce how we build and extract these feature sets in detail.

#### 2.2.1 Clinical-related features

The clinical-related feature set encompasses essential depression and anxiety indicators extracted from EMRs with comprehensive descriptions (shown in Appendix Table A1). This feature set was developed through a collaborative approach involving both psychiatrists and LLM analysis. Firstly, the process began with extracting 218 clinical entities from three sections in the EMR system: chief complaint, personal medical history, and psychiatric examination. These entities represent predefined features within the documentation framework of the SMHC EMR system based on psychiatric diagnostic systems, textbooks, and experts’ opinions. Then, we included a supplementary of 44 additional symptoms identified through clinical expertise and diagnostic criteria (e.g., DSM-5 and ICD-10) suggested by psychiatrists. We then utilized the Gemini 1.5 Pro [39] to generate descriptions for all clinical entities, using the Chinese version of the DSM-5 guidance [3] as a reference, leveraging the model’s strong extended context window capability. Through iterative psychiatric review, redundant and irrelevant items specific to depression and anxiety were eliminated, resulting in a refined set of 138 validated clinical-related features.

After rigorously defining the clinical-related features, we leverage LLM to judge whether these symptoms occur in the diagnostic conversation. In this study, we employed Qwen2-72B-Instruct [47] as the base model to extract clinical and assessment-related features from the clinical dialogue, due to its proficiency in processing the Chinese language and suitability for offline deployment in hospital settings. SFT is a technique that adapts large language models to downstream tasks through supervised learning on domain-specific data. To better adapt to the specific medical terminology and clinical reasoning patterns in our healthcare context, we fine-tuned the model using psychiatrists’ annotations in EMRs. We present the prompt used to generate clinical-related features and fine-tune LLM in Table 1. The fine-tuning was implemented using LLaMA-Factory^1^, and the inference process utilized vLLM^2^. The experiment was conducted on 4 A100 GPUs.

**Table 1:**
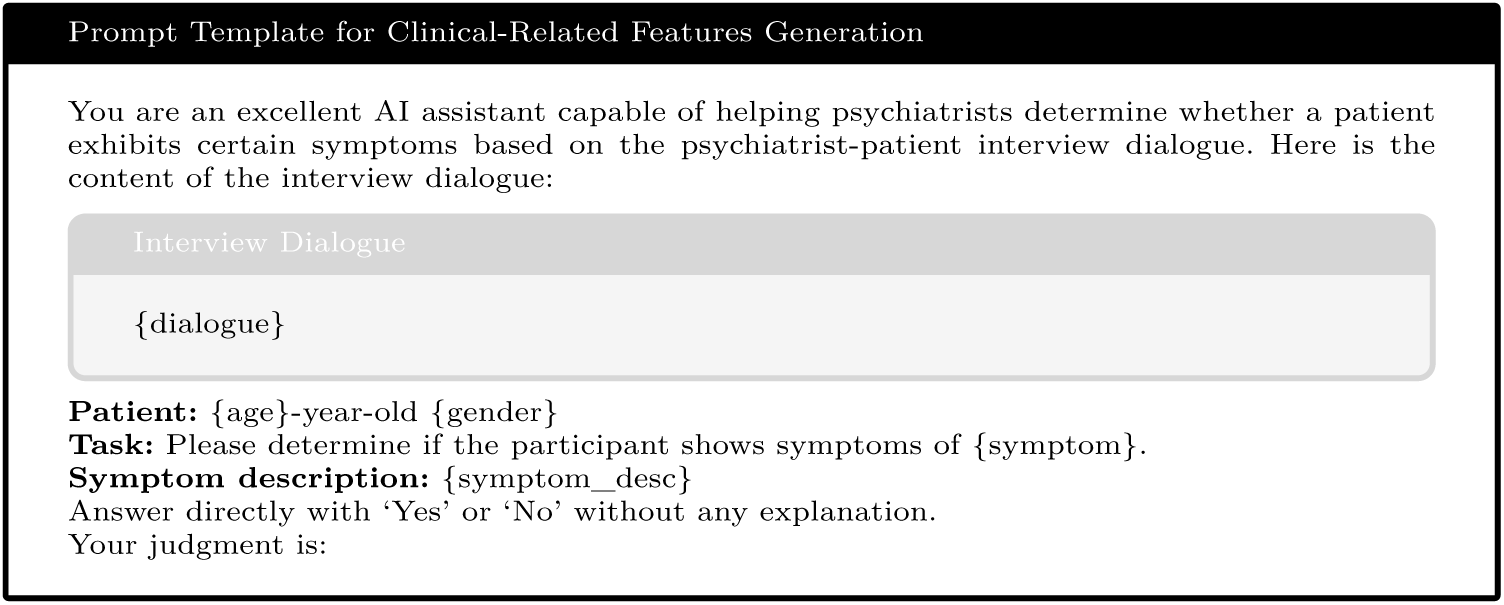
Prompt template for clinical-related feature generation. The content within the curly braces is the demographic, symptom descriptions, and dialogue information that form the prompt.

We first began with structuring EMR data to create reliable training labels for the SFT. Since EMRs contain unstructured text fields where psychiatrists document patient information, we employed the LLM to analyze these 1,160 EMRs. For each EMR, we leveraged LLM to evaluate the presence of above mentioned 138 predefined clinical features, including similar expressions and synonyms, generating a boolean value list (yes/no) for each record. The prompt for querying the LLM to generate labels from EMRs is shown in Appendix Table A3.

Secondly, we implemented a rigorous filtering process for choosing high-quality data for SFT. We first leverage LLM to verify whether the information recorded in EMRs was adequately reflected in the interview dialogue transcripts, yielding 877 valid examples. Then, we collaborated with specialist psychiatrists to establish comprehensive evaluation criteria, encompassing five standards for psychiatric examination, one for chief complaints, and five for present illness history assessment. By using these criteria as the prompt (shown in Appendix Table A3), we employed the LLM to evaluate each case and select the top 60% (527 examples) as high-quality cases based on the total score. From these high-quality cases, we allocated 477 cases for the SFT and 50 cases for the high-quality test set. The 50 high-quality test cases and 633 lower-quality cases are combined as a completed test set to evaluate the accuracy of clinical-related feature extraction.

Subsequently, we fine-tuned the Qwen2-72B-Instruct model with Low-Rank Adaptation (LoRA) [19]. The LLM SFT involves training a pre-trained model on datasets with explicit input-output pairs to optimize the model’s performance on specific down-stream tasks. LoRA is a parameter-efficient fine-tuning technique that adds small, trainable rank decomposition matrices to the LLM’s existing weights, allowing for efficient model adaptation while keeping most of the original model parameters frozen. The model was trained using the following hyperparameters: LoRA rank of 8, LoRA alpha of 16, batch size of 8, and an initial learning rate of 1e-4 for 7,000 steps. During inference using the vLLM framework, we restricted the model’s output to a single token “Yes” or “No” as the binary output, while we also extracted the probability output for these two tokens from the whole vocabulary. After normalization of the probabilities, along with the binary outputs, we formed 276 features in the clinical-related feature set.

#### 2.2.2 Assessment-related features

The assessment-related feature set incorporates data from six validated psychiatric rating scales, combining self-rating and peer-rating instruments. Self-rating scales include SCL-90 [9], SDS [49], and SAS [48], while peer-rating scales comprise HAMD [15], HAMA [16], and MADRS [43], totaling 177 items in all. These scales were selected for their proven reliability in clinical practice and research, offering comprehensive symptom coverage.

We designed two meta-prompts to enable the LLM to mimic both psychiatrists and patients in rating assessment scales in a zero-shot manner, as illustrated in Appendix Table A2. The scales’ content and rating guidelines were integrated into the prompts for LLM to generate the features. For instance, when extracting features related to the first item of the HAMD, which measures depressed mood, we use the peer-rating meta-prompt to instruct the LLM to evaluate the severity of the patient’s depressed mood on a 0-4 scale based on age, gender, and conversation content, where 0 indicates the absence of depression and 4 represents severe depression. When the conversation lacks sufficient information about the depressed mood, the LLM is prompted to return “NULL”. Similar to the clinical-related feature extraction, we extracted and normalized the logits of related tokens from the last layer of LLM and served as the features for classification and prediction tasks, resulting in a total of 1,199 features. We did not SFT the LLM for assessment-related feature extraction, since we do not have sufficient assessment scale labels.

#### 2.2.3 Linguistic features

In addition to the features generated by LLM, we extracted verbal features through two bag-of-words approaches: LIWC [33] and TF-IDF [36], both of which measure the frequency of word occurrence within a document. The LIWC tool is specifically designed to provide rich insights into psychological states, including emotions, thinking styles, and social concerns. It comprises word counts for 63 categories, including 52 categories related to linguistic counts (e.g., function words, common verbs, numbers, etc.), psychological processes (e.g., affect, sociality, cognition, perception, drive, etc.), and personal concern (e.g., work, home, religion, etc.), as well as 7 emotional categories (e.g., happy, sad, fear, etc.) and 4 general text metrics (e.g., the number of unique words, words in LIWC dictionary, etc.). We normalized the LIWC category counts by the total number of words.

The TF-IDF algorithm, which stands for Term Frequency-Inverse Document Frequency, is a popular technique used in text analysis to determine the importance of words within a document or collection of documents. Unlike simple word counting, TF-IDF considers both how often a word appears in a specific document and how common or rare that word is across all documents. This approach helps identify words that are particularly characteristic or important to specific documents. In this study, TF-IDF was applied alongside LIWC to provide a more comprehensive analysis of the verbal features in the documents, offering insights into both the frequency and relevance of words used by the subjects. We applied Jieba^3^ for Chinese character segmentation, resulting in a total of 27,000 features.

#### 2.2.4 Acoustic features

In addition to examining the verbal aspects of participants’ speech, we preprocessed the audio and extracted low-level acoustic and prosodic features using the OpenSMILE toolkits [12]. The audio recordings were manually edited to obscure names, addresses, and personally identifiable information before analysis. To reduce the impact of environmental noise and the varying distance from the microphone to the participant on recording quality, we used the pyAudacity toolkit ^4^ and the FFmpeg-normalized toolkit^5^ to reduce the noise with a parameter of 12dB and normalize the volume to -23dB respectively. OpenSMILE is a versatile, customizable suite of acoustic features for signal processing and machine learning applications. We utilized OpenSMILE’s emobase_live4 configuration to extract the following LLDs: intensity, loudness, 12 MFCCs, pitch (F0), voicing probability (VoiceProb), F0 envelope (F0env), 8 line spectral frequencies (LSF), and Zero-Crossing Rate (ZCR). Next, we applied various functions to these LLDs and their delta coefficients, including minimum and maximum values with their relative positions (minPos and maxPos), range, mean, linear regression coefficients (linregc1–2), linear and quadratic error, standard deviation (STD), skewness, kurtosis, quartile values (quartile1–3), and interquartile ranges (iqr1-2, iqr2-3, iqr1–3). This process yielded 988 features to represent each speech utterance. Before LLD computation, pauses and silences were eliminated from the speech to create a continuous signal. We then extracted 988 emotion-based prosodic features using a 100 ms sliding window over the entire speech sample. Lastly, we calculated these emotion-based features’ maximum, minimum, mean, and standard deviation to compose the final set of OpenSMILE features, totaling 3,952 features.

### 2.3 Classification method

As explained in previous sections, we extracted five feature sets using LLM and existing toolkits: clinical-related, assessment-related, LIWC, TF-IDF, and OpenSMILE features. Subsequently, we built a machine learning pipeline to fuse the outputs from multiple feature sets to predict the appearance of the symptom and classify diagnostic groups, which was implemented using Scikit-learn 1.2.0 in Python 3.10. Notably, as detailed in Section 2.2.1, we fine-tuned the LLM using 138 high-quality clinical annotations to improve its ability to identify clinical concepts. We excluded diagnostic labels from this process to prevent data leakage.

To ensure robust validation, we employed 10-fold cross-validation (10-fold CV). This method involves dividing the data into 10 subsets, iteratively training the model on 9 subsets, and testing it on the remaining subset. The process is repeated 10 times, with each subset serving as the test set once, and the model’s performance is averaged across all iterations. To address the challenge of class imbalance, we applied the Synthetic Minority Oversampling Technique (SMOTE) [5], which generates synthetic data for minority classes. Furthermore, we performed z-score standardization on all features, resulting in standardized features with a mean of 0 and a standard deviation of 1. This step ensures that all features are on a comparable scale, preventing any single feature from dominating the analysis due to its magnitude.

We also implemented probability calibration to standardize predictions from each feature set. This process involved an internal CV on the training set of the outer CV to obtain the probability distribution on training data, which were then used to calibrate test set predictions [46]. Moreover, based on the feature importance ranked by the classifier on training data, we filtered out features whose importance values fell below the mean to reduce unimportant features. For the final prediction, we employed a late fusion technique, a multi-modal machine learning approach that involved averaging the standardized prediction outputs from all feature sets to produce the final output. This method allows for the integration of diverse information sources while maintaining the integrity of each feature set’s contribution to the final prediction.

### 2.4 Performance metrics

To evaluate the performance of the LLM in extracting clinical features from interview dialogues, we employed standard information extraction metrics: precision and recall. Precision measures the proportion of correctly identified symptoms among all symptoms extracted by the LLM, while recall measures the proportion of symptoms correctly extracted from the EMR annotations. Given that psychiatrists may not document every symptom mentioned during interviews in the EMRs, recall serves as a particularly valuable metric in our evaluation framework. Precision and recall are calculated as follows: Precision = TP / (TP + FP); Recall = TP / (TP + FN), where TP (True Positives) represents symptoms correctly identified by both the LLM, FP (False Positives) represents symptoms incorrectly extracted by the LLM, and FN (False Negatives) represents symptoms present in the EMR but missed by the LLM.

For classification and prediction tasks, we utilize a comprehensive set of standard metrics. Our analysis primarily focuses on balanced accuracy (BAC), which is particularly effective for imbalanced datasets by averaging sensitivity (SEN) and specificity (SPE). This metric provides a robust measure of overall performance, accounting for both true positive and true negative rates. In addition to BAC, we employ several other metrics to ensure a thorough assessment: positive predictive value (PPV), negative predictive value (NPV), and area under the precision-recall curve (AUPRC). The AUPRC, like BAC, is well-suited for machine learning tasks involving imbalanced data and offers valuable insights into model performance across various classification thresholds.

Understanding the key distinguishing features among various mental health conditions is crucial for improving diagnostic accuracy, developing targeted interventions, and enhancing our overall comprehension of these disorders. To address this critical need, we employed a comprehensive approach to identify the most important features distinguishing between different mental health conditions. We utilized various feature sets, including LLM-generated clinical and assessment-related features, LIWC categories, and TF-IDF terms, and applied the Mann-Whitney U test with FDR correction across all feature sets to calculate p-values and measure feature importance. Features are ranked by their p-values, with those below 0.05 indicating a statistically significant difference between the two groups.

## 3 Results

### 3.1 Sample

The study included 1,160 individuals, yielding about 15,000 minutes of speech data. All participants received diagnoses based on the ICD-10 [20]. The sample comprised 553 participants diagnosed with “Depressive Episode” or “Depressive Disorder” (DP), 426 diagnosed with “Anxiety Disorder” or “Anxiety State” (ANX), and 181 classified as “Others” (patients not diagnosed with DP or ANX). Table 2 presents the demographic characteristics of the participants. Moreover, based on the clinical annotations of symptom episodes in the EMRs, we categorized the participants into four groups: patients who experienced/presented anxiety symptoms (A), participants who experienced/presented depressive symptoms (D), participants who experienced/presented mixed depressive and anxiety symptoms (M), and participants without experienced/presented depressive and anxiety symptoms (N).

**Table 2:** Demographics of all participants.

|  | DP (N = 553) | ANX (N = 426) | Others (N = 181) |
| --- | --- | --- | --- |
| **Age** | 29.2 ± 10.0 | 34.0 ± 12.0 | 27.5 ± 11.1 |
| **Gender** |  |  |  |
| **Female** | 377 (68.2%) | 288 (67.6%) | 104 (57.5%) |
| **Male** | 176 (31.8%) | 138 (32.4%) | 77 (42.5%) |
| **Occupation** |  |  |  |
| **Employed** | 266 (48.1%) | 250 (58.7%) | 74 (40.9%) |
| **Student** | 201 (36.3%) | 98 (23.0%) | 81 (44.8%) |
| **Unemployed** | 46 (8.3%) | 44 (10.3%) | 14 (7.7%) |
| **Unknown** | 23 (4.2%) | 15 (3.5%) | 7 (3.9%) |
| **Retired** | 9 (1.6%) | 19 (4.5%) | 4 (2.2%) |
| **Dropped out** | 8 (1.4%) | NaN | 1 (0.6%) |
| **Personality** |  |  |  |
| **Introvert** | 274 (49.5%) | 198 (46.5%) | 106 (58.6%) |
| **Extrovert** | 148 (26.8%) | 114 (26.8%) | 34 (18.8%) |
| **Gentle** | 46 (8.3%) | 38 (8.9%) | 13 (7.2%) |
| **Sensitive** | 29 (5.2%) | 22 (5.2%) | 4 (2.2%) |
| **Strong welling** | 12 (2.2%) | 14 (3.3%) | 8 (4.4%) |
| **Others** | 44 (8.0%) | 40 (9.4%) | 16 (8.8%) |

### 3.2 LLM-generated clinical-related features evaluation

We evaluated the performance of LLM-generated clinical symptoms on the entire test samples and those with high-quality EMR, as shown in Table 3. Our evaluation of LLM-based clinical symptom extraction demonstrated a significant performance improvement after the SFT, with the accuracy increased from 81.2% to 86.9% on the test set and 83.7% to 89.1% on the high-quality test set. The recall metric showed substantial improvements, increasing from 66.1% to 81.1% on the whole test set and from 74.0% to 86.1% on the high-quality test set, indicating enhanced capability in identifying symptoms documented by psychiatrists in the EMR. Meanwhile, precision improved from 81.2% to 87.4% on the test set and from 84.2% to 89.5% on the high-quality test set. This precision increase, coupled with recall improvement, suggests that the fine-tuned model became more comprehensive in detecting symptoms from clinical dialogues.

**Table 3:** Performance comparison of LLM-generated clinical-related features between Zero-shot and SFT approaches.

|  | Test set |  |  | High-quality test set |  |  |
| --- | --- | --- | --- | --- | --- | --- |
|  | Precision | Recall | Accuracy | Precision | Recall | Accuracy |
| Zero-shot | 81.2% | 66.1% | 81.2% | 84.2% | 74.0% | 83.7% |
| the SFT | 87.4% | 81.1% | 86.9% | 89.5% | 86.1% | 89.1% |

We present a comparative analysis of classification performance using clinical-related features extracted by the LLM in Figure 2, comparing three feature sets: features extracted in a zero-shot manner, features extracted from the fine-tuned LLM, and psychiatrists’ annotations derived from EMRs. Across all classification tasks, features from the fine-tuned LLM consistently demonstrate superior performance. For instance, in distinguishing between depression and anxiety diagnoses (A vs. D), the fine-tuned LLM achieves a BAC of 74.8%. In identifying depression (D vs. N) and anxiety symptoms (A vs. N), the BAC reaches 79.8% and 72.2% respectively. These results underscore the potential of fine-tuned LLMs for accurate and automated clinical manifestation extraction.

**Fig. 2:**
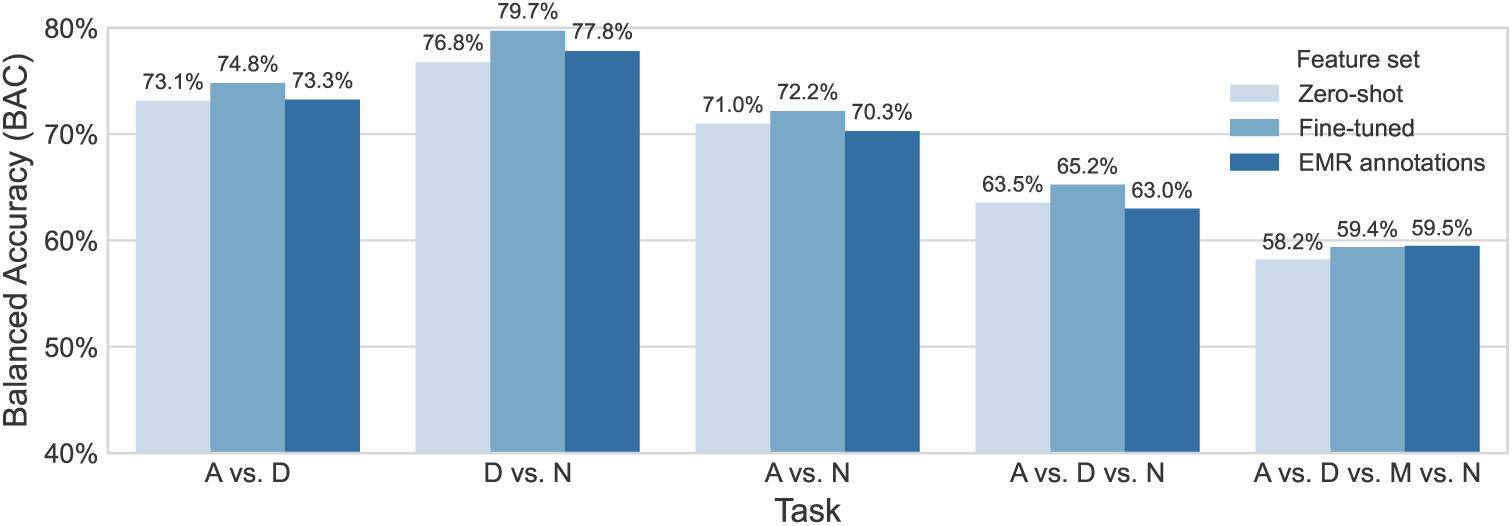
Comparative analysis of classification performance using the clinical-related features extracted by LLM in zero-shot, the SFT, and the annotations from EMRs across different classification tasks.

### 3.3 Classification of diagnostic groups

The results of automated classification tasks for distinguishing between ANX, DP, and Others groups (not diagnosed with ANX or DP) using various linguistic and LLM-generated features are shown in Table 4. For the binary classification task (ANX vs. DP), the model achieved a BAC of 75.5%, an F1 score of 0.762, and an AUPRC of 0.824, indicating good overall performance (permutation test p-value < 0.01, same for other tasks). In the three-way classification task (ANX vs. DP vs. Other), the model’s performance was achieved with a BAC of 65.6% and an F1 score of 0.656, presenting a significant gain compared to the majority baseline (47.7%).

**Table 4:**
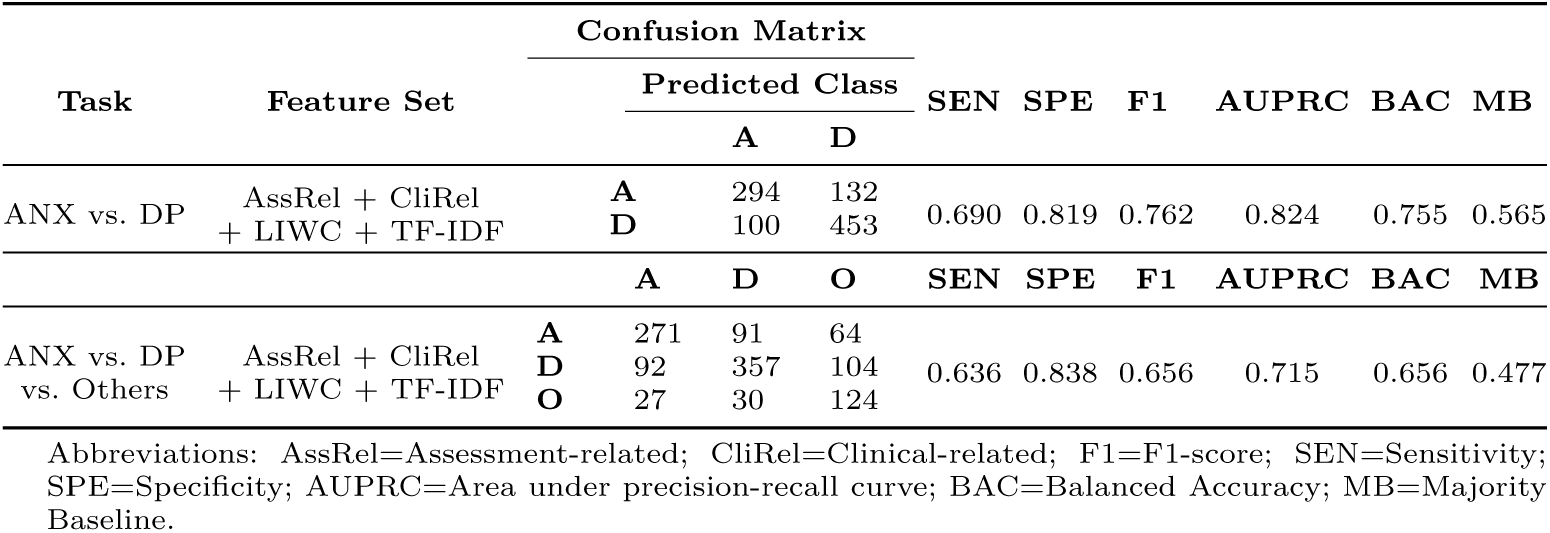
Results for classification of ANX, DP, and Others groups.

### 3.4 Prediction of depression and anxiety symptoms

In addition to identifying diagnostic results by ICD-10 code, we predicted whether participants exhibited symptoms of depression, anxiety, mixed depression/anxiety, or no symptoms at all, as shown in Table 5. In the anxiety vs. no anxiety (A vs. N) classification task, the model achieved a sensitivity of 0.683 and specificity of 0.810 for detecting anxiety, with an overall F1 score of 0.754 and BAC of 74.7%. For the depression vs. no depression (D vs. N) task, the model performed slightly better, with a sensitivity of 0.806 and specificity of 0.737 for detecting depression, resulting in an F1 score of 0.783 and a BAC of 77.2%. When distinguishing between anxiety, depression, mixed symptoms, and no depression and anxiety symptoms (A vs. D vs. M vs. N), we achieved an AUPRC of 0.606 and a BAC of 60.7%, which achieved a significant improvement of about 30% compared to the majority baseline.

**Table 5:**
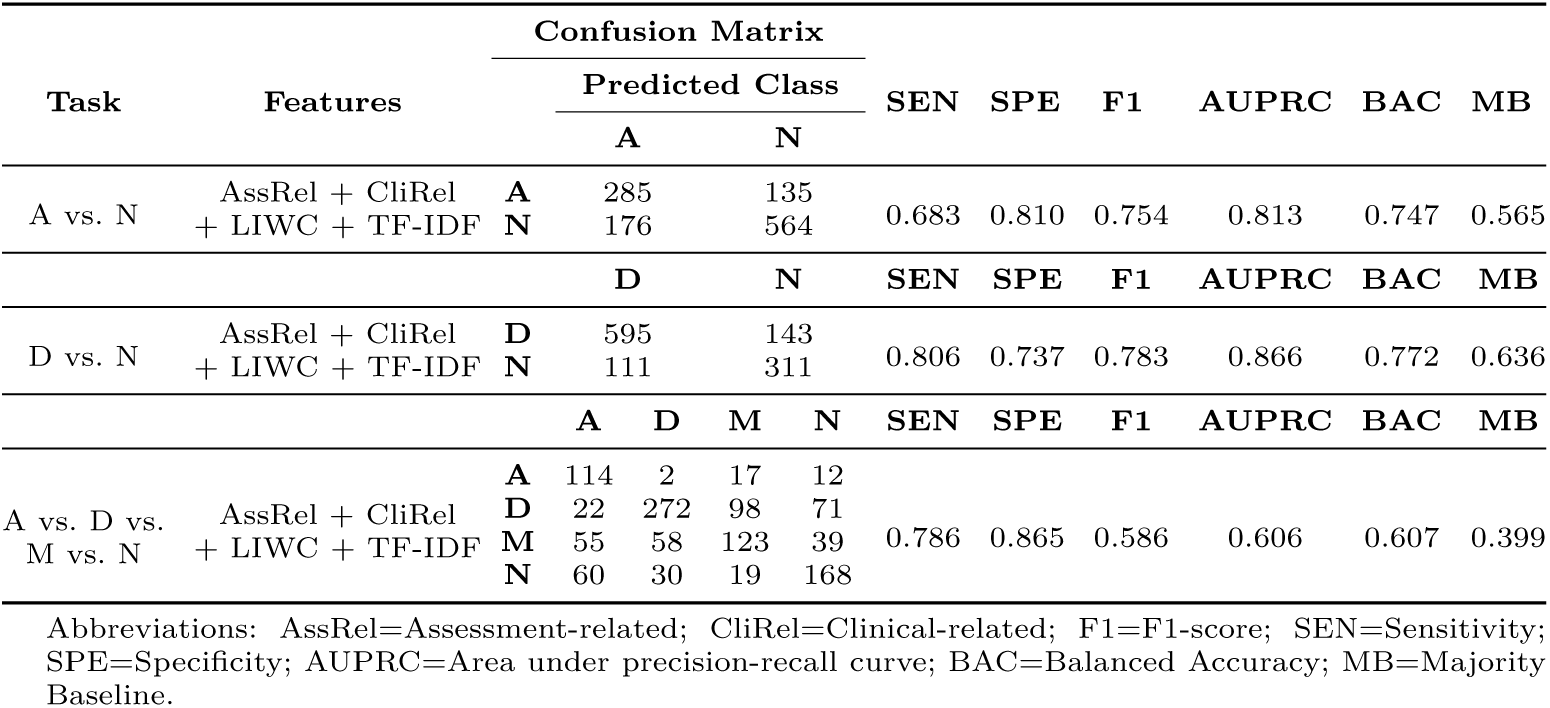
Results for classification of participants with depression (D), anxiety (A), mixed depression and anxiety (M), and no depression and anxiety symptoms (N).

### 3.5 Interpretability

The analysis revealed distinctive patterns across different mental health conditions and feature sets (Table 6). In differentiating ANX from DP, clinical-related features emphasized anxiety-specific symptoms such as “Unable to relax”, “Uncontrollable restlessness”, and “Anxiety”, contrasting with depressive symptoms like “Sadness” and “Anhedonia”. Assessment measures showed a mixed profile, with both anxiety indicators (HAMD_Somatic anxiety) and depression markers (HAMD_Depressed mood). LIWC analysis revealed heightened use of anxiety and fear-related language, and TF-IDF identified anxiety-related terms. For depression detection, clinical-related features highlighted core depressive symptoms, with “Depressed mood”, “Loss of interest”, and “Anhedonia” emerging as primary indicators of depression. The assessment-related features showed strong signals from SCL-90 scales, particularly in items related to feelings of sadness and loss of interest. LIWC analysis identified significant usage patterns in sadness-related words and negative emotions, while TF-IDF analysis captured depression-specific terms and notably, negation patterns (e.g., “Don’t want”, “No”, etc.). For anxiety identification, clinical-related features strongly centered on anxiety manifestations, such as “Unable to relax”, “Anxiety”, and “Worry.” The assessment-related features prominently featured inner tension and somatic anxiety, along with various SCL-90 anxiety-related items. Both LIWC and TF-IDF analyses consistently identified anxiety-specific language patterns, with LIWC showing “Anxiety” and “Fear” as top features, and TF-IDF highlighting terms related to physical symptoms (e.g., “Palpitations”, “Heartbeat”, etc.) and worry.

**Table 6:**
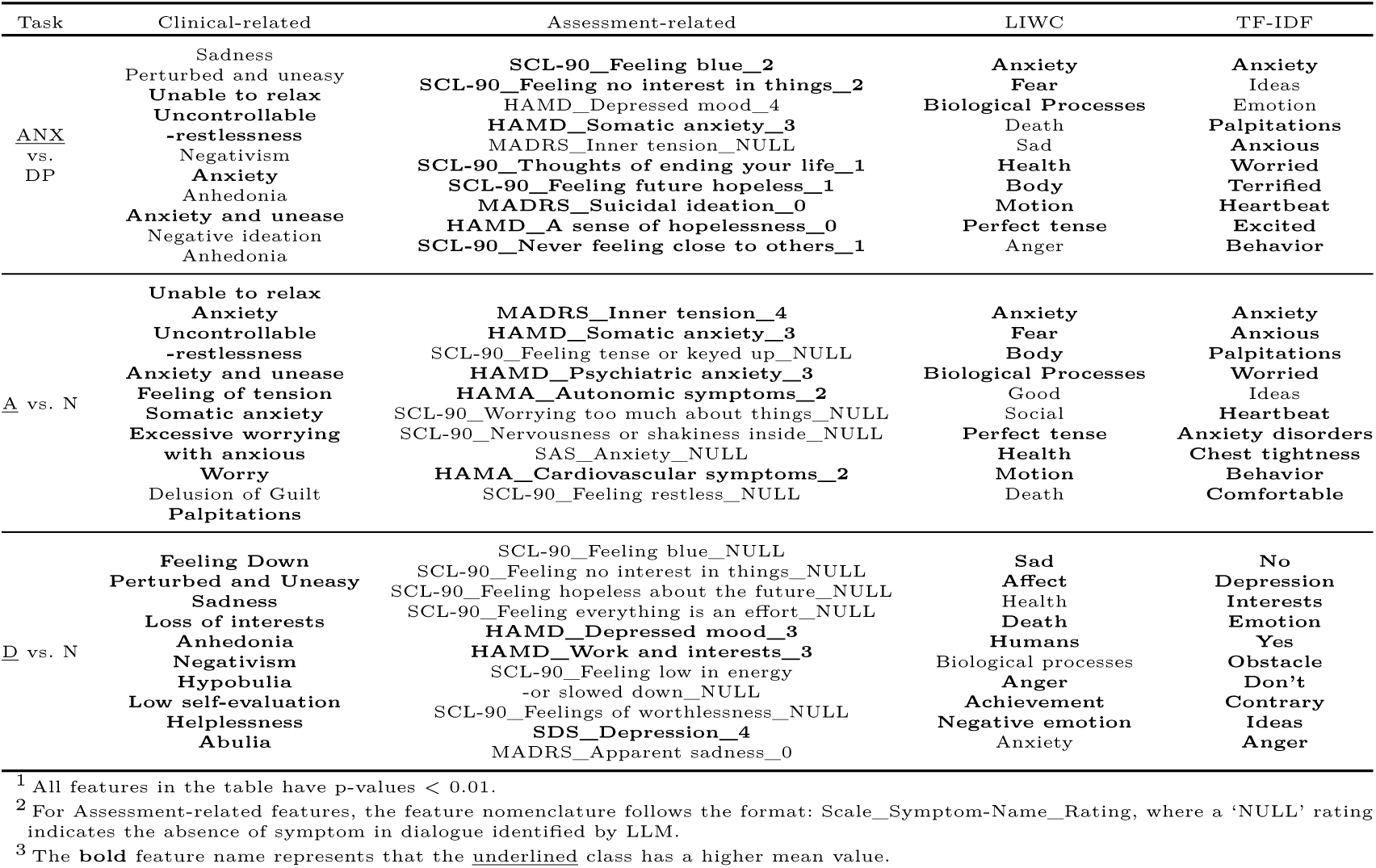
Top ten salient features for each feature set in paired classification tasks.

## 4 Discussion

Inspired by promising early research on digital phenotypes for diagnosing and classifying symptoms in psychiatric patients, we investigated using signal processing and state-of-the-art LLM to capture symptom-related expression cues in outpatient conversations. Subsequently, we developed an ensemble classification pipeline to automatically differentiate between clinical diagnostic outcomes and the presence of symptoms.

Although recent studies have demonstrated promising capabilities of utilizing LLMs in medical diagnosis [14], applications in mental health have predominantly centered on developing conversational agents [38], while the potential of extracting precise symptoms from psychiatric conversations for evidence-based diagnosis has not been fully explored. In this study, we investigated the efficacy of LLM in detecting clinical and assessment-related symptoms. Our investigation revealed that without any additional training, the model achieved a recall rate of 77.3% on high-quality dialogue-case pairs, and increased to 86.1% by fine-tuning the LLM using clinical annotations. This aligns with recent observations regarding LLMs’ strong zero-shot performance in healthcare domains and the fine-tuning could further boost LLM performance [24]. Furthermore, this enhanced base capability led to substantial improvements across all downstream classification and prediction tasks (e.g., the classification accuracy for ANX and DP increased from 72% to 75%). Current approaches to automated symptom detection predominantly rely on traditional natural language processing methods with predefined linguistic categories or rule-based systems [45, 46], which often struggle to capture the complex presentation of psychiatric symptoms in natural conversation. Some researchers have explored the use of LLMs to assist in medical information retrieval [26]. We further investigated the information extraction capabilities in clinical dialogues and enhanced them through SFT.

Our study demonstrated moderate to high performance in anxiety symptom detection (BAC=74.7%, AUPRC=0.813), depression symptoms detection (BAC=77.2%, AUPRC=0.866), and a four-class classification of patients with anxiety/depression/mixed/none symptoms (BAC=60.7%, AUPRC=0.606). As shown in the summarization of existing literature (in Appendix A4), while anxiety detection in social media text has demonstrated promising results with high accuracy [35], the performance of similar methods on spoken language data, such as interview transcripts and therapy dialogues, remains limited with accuracy rates below 65%. Recent advances combining LLM embedding with acoustic features have shown improved results, reaching 75% accuracy in a small cohort of 65 patients [21]. While depression detection studies have reported wide-ranging accuracy rates (65%-95%), some results should be interpreted with caution due to several methodological limitations: small sample sizes [45], reliance on PHQ screening tools rather than clinical diagnoses [17], data collection in structured experimental settings [10], and not studied the first-episode outpatients in real-world, unstructured clinical environments. Our study leverages clinical diagnoses from psychiatrists and achieved a moderate to high accuracy in this more challenging and unstructured clinical setting demonstrating the robustness of our approach. This success particularly highlights the potential of LLMs in extracting and analyzing clinical symptoms for predicting anxiety and depression in outpatient populations, offering a more ecologically valid and scalable solution for mental health screening and monitoring.

DP and ANX present significant diagnostic challenges due to their high prevalence, frequent comorbidity, and overlapping symptomatology [18]. By leveraging LLM-generated features, our approach achieved robust performance in distinguishing these disorders, with a BAC of 75.5% and AUPRC of 0.824 for binary classification between DP and ANX, and the performance outperformed the directly using LLMs as classifiers (see Appendix A5). In the more challenging multi-class scenario (ANX vs. DP vs. Others), the model maintained reasonable performance with a BAC of 65.6% and AUPRC of 0.715. Prior approaches to differentiating depression and anxiety disorders, such as cognitive tasks [34] and structured questionnaires [28], have achieved accuracy rates of 70-80%. In addition, we tested the classification performance of each assessment scale as the feature set, where the results are presented in Appendix Table A4. We observed that assessment-related features, particularly from scales like SCL- 90, HAMD, and MADRS, showed strong discriminatory power across all comparisons, and early fusion and late fusion present similar classification performance. A potential reason is that these scales contain sufficient depression-related symptoms, which are key components for differentiating different groups. To our knowledge, no study has explored the objective diagnosis of DP and ANX using speech data from clinical interviews, potentially due to a lack of data and inherent subjectivity. Our study addresses a critical gap by analyzing the linguistic and symptom-related markers in various participant groups, providing objective cues to assist psychiatrists.

The feature analysis provides several key insights into the differential characteristics of different groups of participants, as shown in Table 6. We also illustrate the distribution of clinical and assessment-related features for each group of participants in Appendix Table A3 and Table A4. The clinical-related features demonstrate clear condition-specific patterns: features that show more importance in patients with depression cluster around mood (sadness and disappointment) and motivational disturbances (anhedonia, reduced volition), while anxiety features predominantly reflect an inability to relax and worry. The observation for depression is in line with previous studies which also observed that patients with depression presented blunted facial affect and increased sadness in language [21, 42] and anhedonia is specific to depression [8]. For anxiety recognition, the consistency of findings across different feature sets strengthens the reliability of these discriminators. For instance, the prominence of somatic symptoms in anxiety, captured in both assessment-related features and TF-IDF terms, suggests this could be a robust marker. Similarly, the persistent appearance of mood-related terms in depression across multiple feature sets reinforces their diagnostic utility.

In summary, this study demonstrates the potential of using LLM to analyze digital biomarkers in speech for automatic assistance in psychosis diagnosis and assessment. Our model achieved promising accuracy in identifying individuals with anxiety and depression symptoms, as well as differentiating between DP and ANX groups. Using LLMs to extract clinically relevant features and rate assessment scales improved the interpretability of the results, offering a novel approach to bridging the gap between automated analysis and clinical practice. While further research is needed, our findings suggest that well-developed LLMs could potentially serve as valuable tools in standardizing psychiatric evaluation and decision-making.

## 5 Limitations

Our study has several limitations that should be addressed in future research. The absence of detailed symptom severity measures during the experiment limits our ability to correlate speech patterns with specific symptom intensities. Additionally, the study’s focus on specific disorders and potential biases in data collection may affect the generalizability of the results. Future work should prioritize the inclusion of comprehensive symptom severity assessments and explore the application of this approach to a broader range of mental health conditions. Besides, in the future, we will collect more data to perform longitudinal analysis, as it could provide insights into how linguistic patterns evolve with symptom progression or treatment response. Furthermore, expanding the use of more advanced LLMs in this context could potentially enhance the extraction of nuanced clinical concepts and provide even more detailed, interpretable insights for clinicians. Validating the model’s performance in diverse clinical settings and with larger, more diverse patient populations will be crucial to ensure its practical utility and generalizability. These advancements could significantly contribute to improving the efficiency and objectivity of consultations for depression, anxiety, and potentially other mental health disorders.

## Data Availability

The data and features processed in the intermediate process in the present study are available upon reasonable request to the authors

## Acknowledgements

This study was funded by 2023-TX-018 from Tianqiao and Chrissy Chen Institute (TCCI) with the Programe of Chen Frontier Lab for AI and Mental Health (TCCI) - Shanghai Mental Health Center (SMHC). J.C. was supported by 82071500 from the National Natural Science Foundation of China and 21XD1423300 from the Program of Shanghai Academic/Technology Research Leader. We deeply appreciate every participant involved in this study and all the efforts made by TCCI and SMHC colleagues who are not on the author list.

## Author contributions

S.X. conceptualized the study, developed the large language model methodology, designed the machine learning pipeline, conducted the experiments, and wrote the original draft. Y.Y. and Y.D. performed clinical concept verification and data analysis. F.L. and S.Z. developed the large language model methodology, performed data processing, and conducted the experiments. T.Y. and H.G. provided technical support and data resources. J.S., X.J., and J.C. supervised the project, acquired funding, and reviewed the final manuscript. N.O. reviewed the final manuscript. Other authors contributed to the clinical data collection. All authors contributed to the manuscript revision and approved the submitted version.

## Data availability

The code and data can be found at https://github.com/Shanda-Group-Ltd/SMHC_llm_psychiatry_study

## A Descriptions of clinical-related features

**Table A1:**
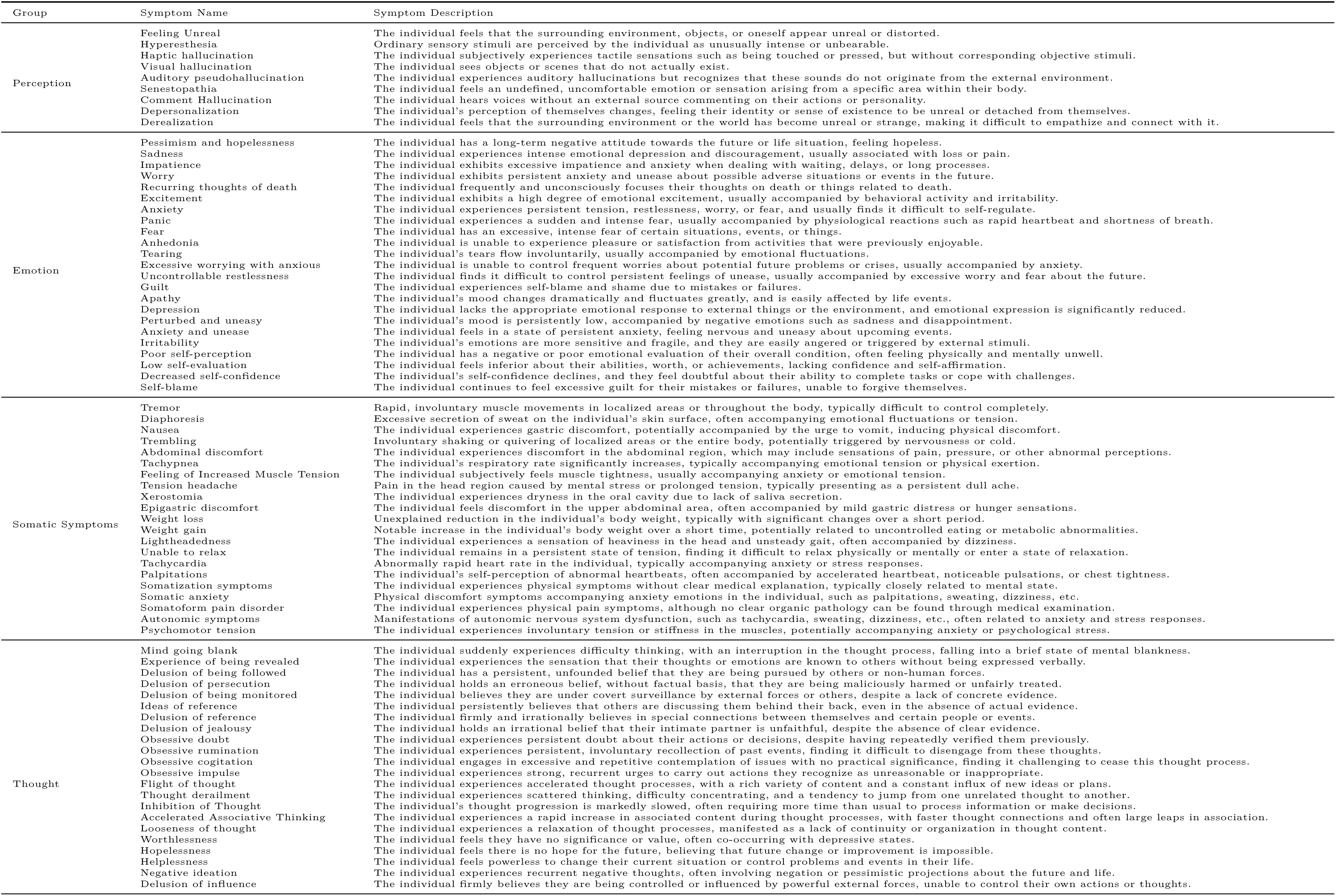
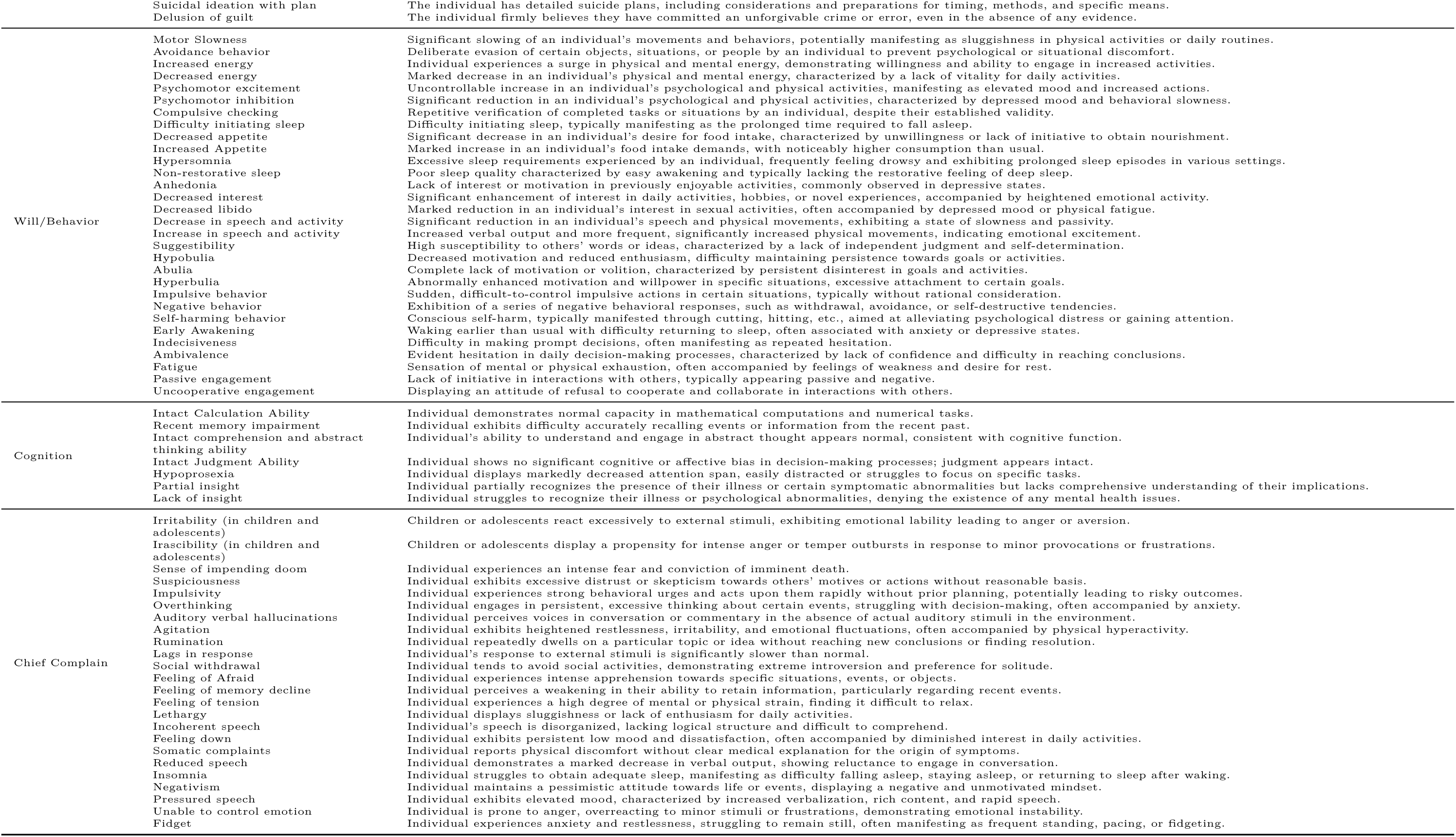
The description of clinical-related features extracted from EMRs and DSM/ICD.

## B Additional classification results

**Fig. A1:**
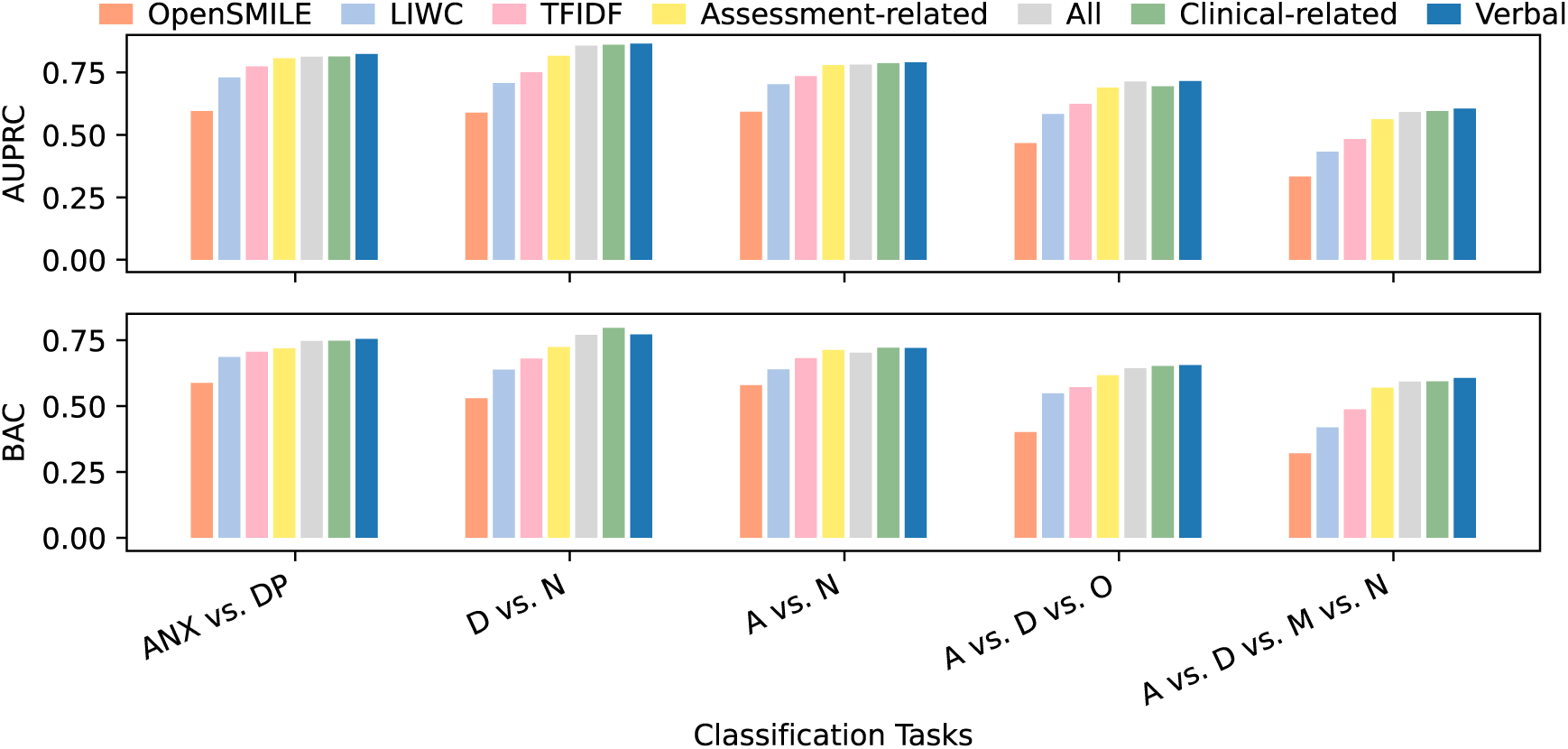
The classification results for different feature sets.

**Fig. A2:**
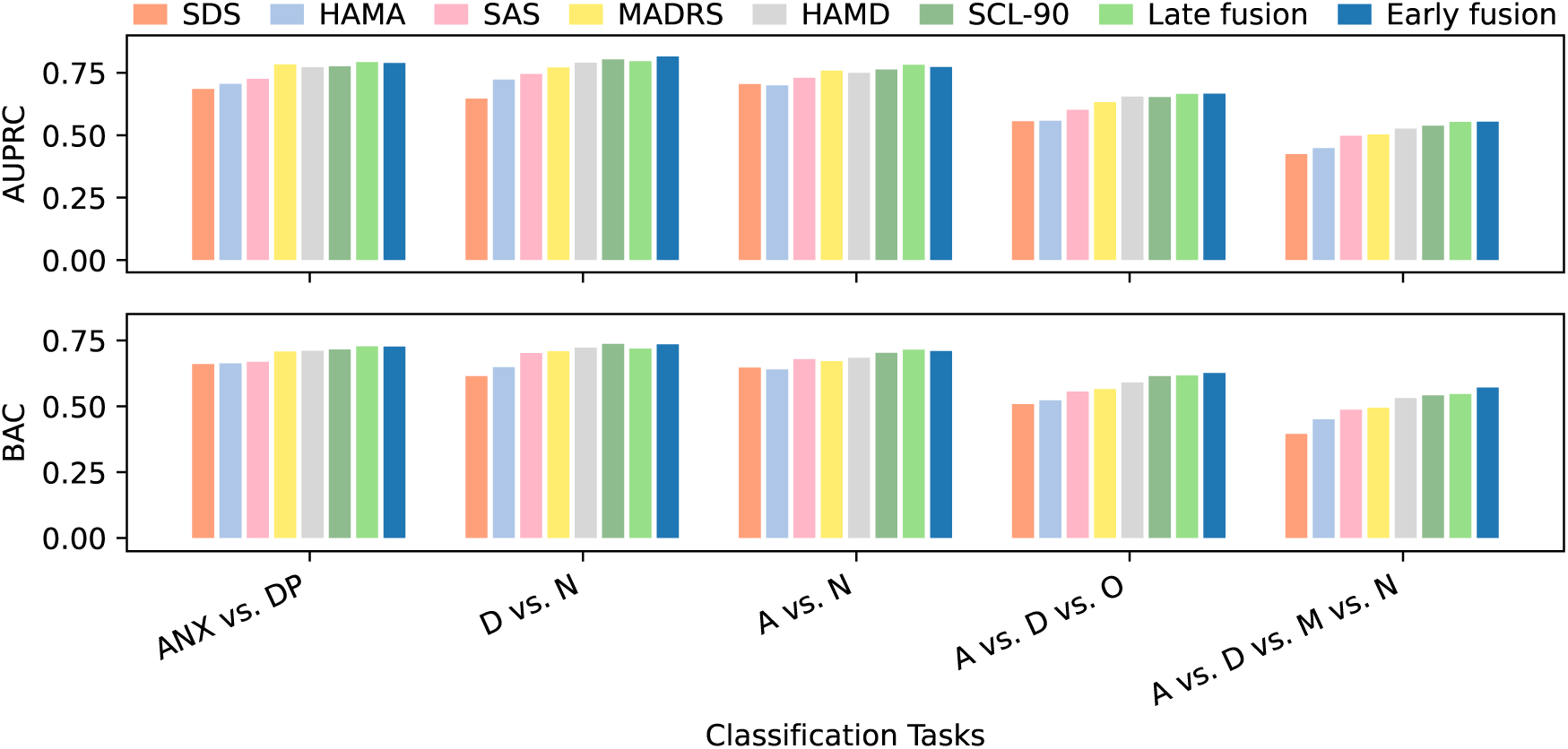
The classification results for different assessment scales. Early fusion combines features before classification, and late fusion merges individual classifier outputs by simply averaging the output probabilities.

## C Prompt Templates

**Table A2:** Prompt templates for assessment-related feature generation.

|  |
| --- |
| Prompt Template for Assessment-Related Features Generation (Peer-Rating Scale)<br>This is a dialogue between a psychiatrist and a patient. Based on the following conversation, please rate the symptom severity according to the {scale} scale, referring to the symptom description and symptom evaluation description:<br>Interview Dialogue<br>{dialogue}<br>**Patient:** {age}-year-old {gender}<br>**Symptom to evaluate:** {symptom}<br>**Symptom description:** {symptom_desc}<br>**Scoring range:** {score_range}<br>**Symptom evaluation description:** {symptom_desc}<br>Please review the dialogue content provided above: determine whether the specified symptom was mentioned in the conversation; If the symptom was not discussed at all, respond with 'null'; If the symptom was indeed mentioned, assign an appropriate score based on the symptom evaluation description provided; Provide only the numerical score without any explanation or reasoning for your choice.<br>The scoring rating is: |
| Prompt Template for Assessment-Related Features Generation (Self-Rating Scale)<br>You are playing the role of a patient who has just finished an interview with a psychiatrist. Your conversation content is as follows:<br>Interview Dialogue<br>{dialogue}<br>The patient you are portraying is a {age}-year-old {gender}.<br>After the conversation, you need to complete a {scale} self-assessment scale.<br>**Symptom to evaluate:** {symptom}<br>**Symptom description:** {symptom_desc}<br>**Scoring range:** {score_range}<br>**Symptom evaluation description:** {score_desc}<br>Please provide your assessment. If this symptom wasn't discussed in your conversation, please answer 'null'. If it was mentioned, please provide an appropriate score based on the description of the symptom evaluation. Give only the score without explaining the reasoning behind it.<br>The scoring rating is: |

**Table A3:**
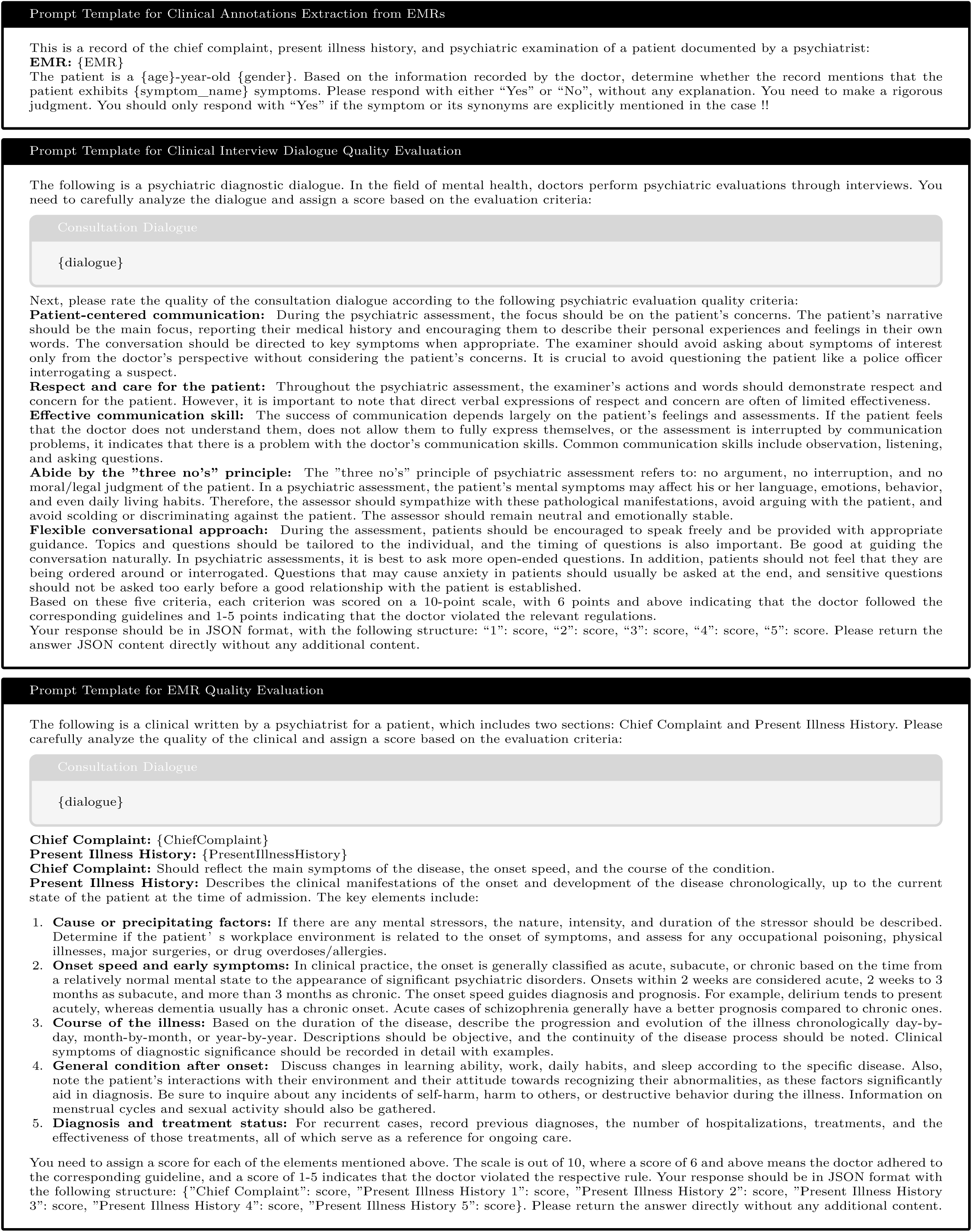
Prompt templates used for the clinical-related annotations generation, dialogue quality evaluation, and EMR quality evaluation.

**Table A4:**
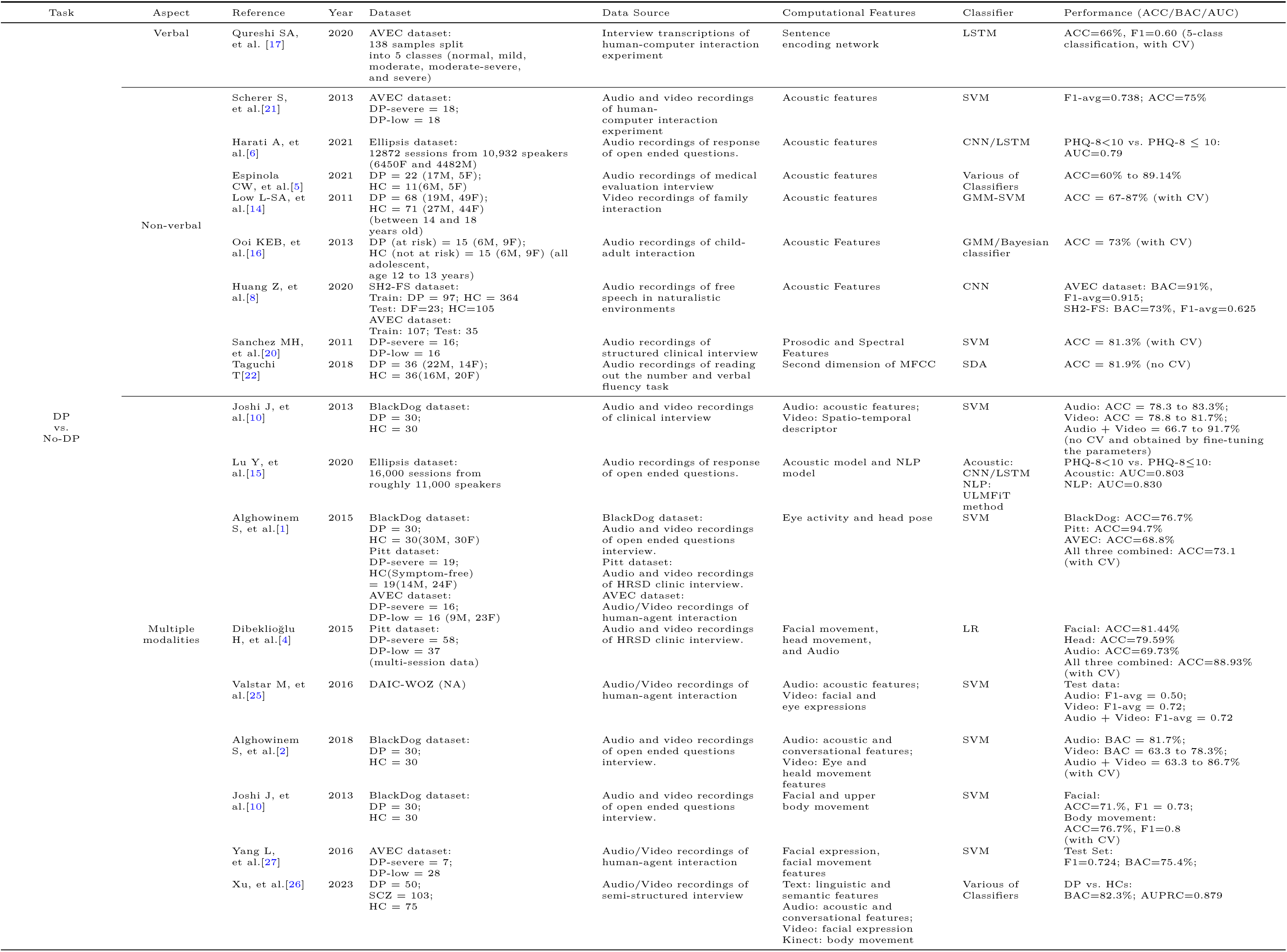
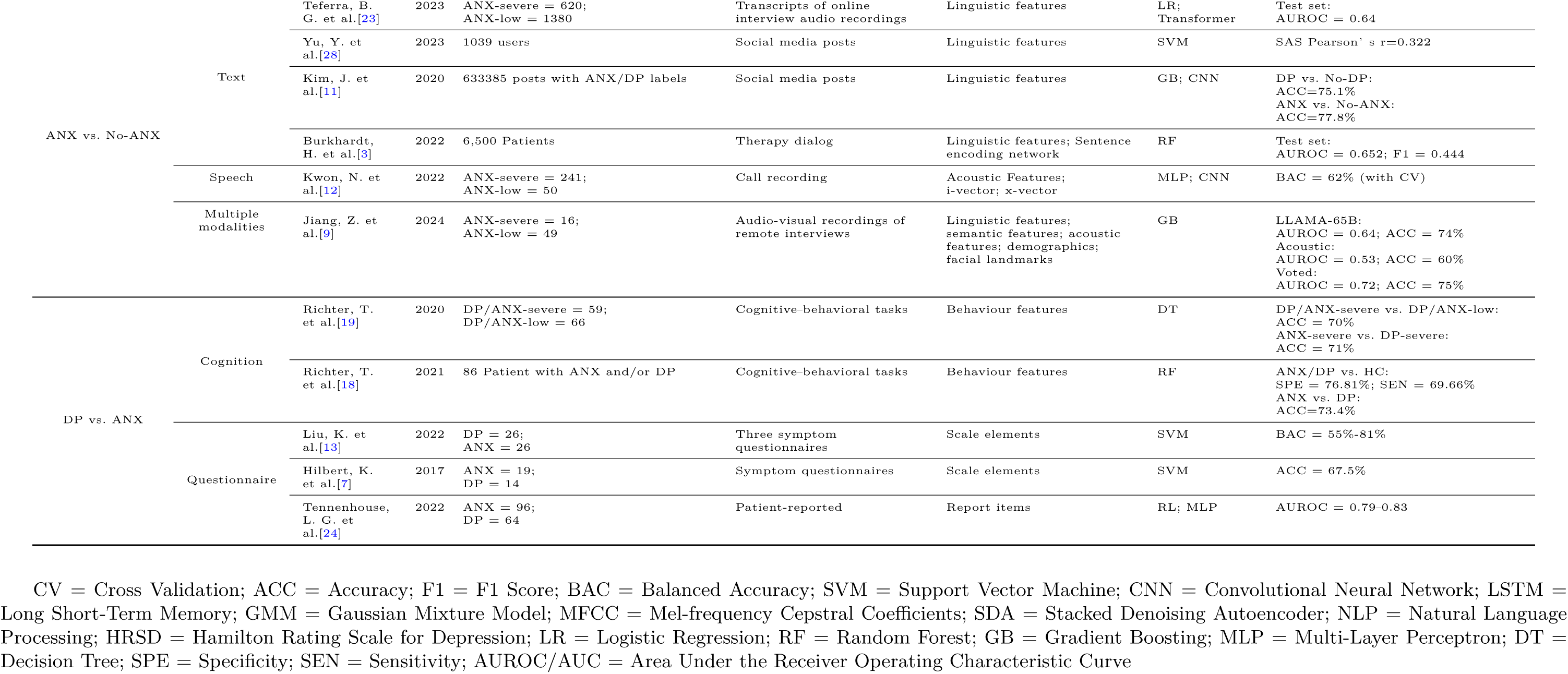
Literature Review of related data-driven studies on identifying depression (DP) and general anxiety disorders (ANX).

## D LLMs as classifiers

We conducted a systematic investigation of diagnostic efficacy in clinical dialogue classification utilizing the Qwen2-72B-Instruct LLM, implementing two distinct methodological approaches: direct classification and intermediate feature extraction. We designed a suitable prompt, added the dialogue content to the prompt to ask the LLM whether it is depression or anxiety, and let the LLM output only one of the two, and then we captured the probabilities of the two judgment output tokens to calculate the classification metrics. To ensure robust evaluation, we employed a stratified data partitioning strategy, allocating 60% of both DP and ANX samples for training, 20% for validation, and 20% for testing. In the SFT paradigm, we fine-tuned the model on the training set with LoRA, employed validation loss as the stopping criterion, and evaluated performance on the held-out test set. For comparative assessment, both zero-shot prompting and intermediate feature extraction approaches were evaluated on the same test set, maintaining consistency across all methods. We observed that using the approach in this paper provides better performance than directly using LLM as the classifier even fine-tuning the model.

**Table A5:**
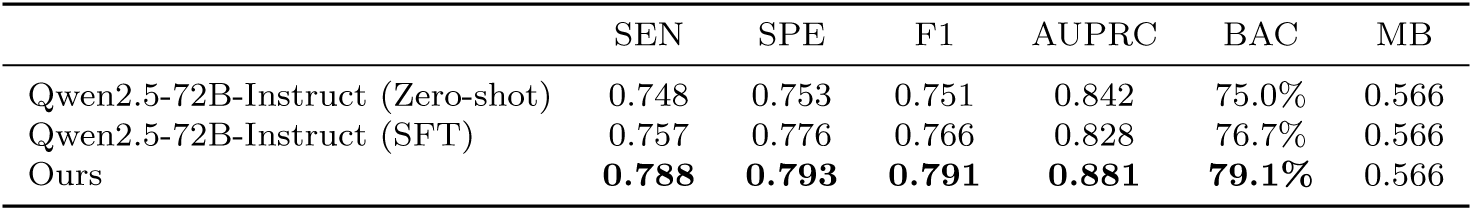
Comparison of classification performance using Qwen2-72B-Instruct (Zero-shot and SFT) and Ours (LLM-generated feature sets with ensemble random forest classifiers) for DP/ANX classification.

## E Distribution illustration of salient features

**Fig. A3:**
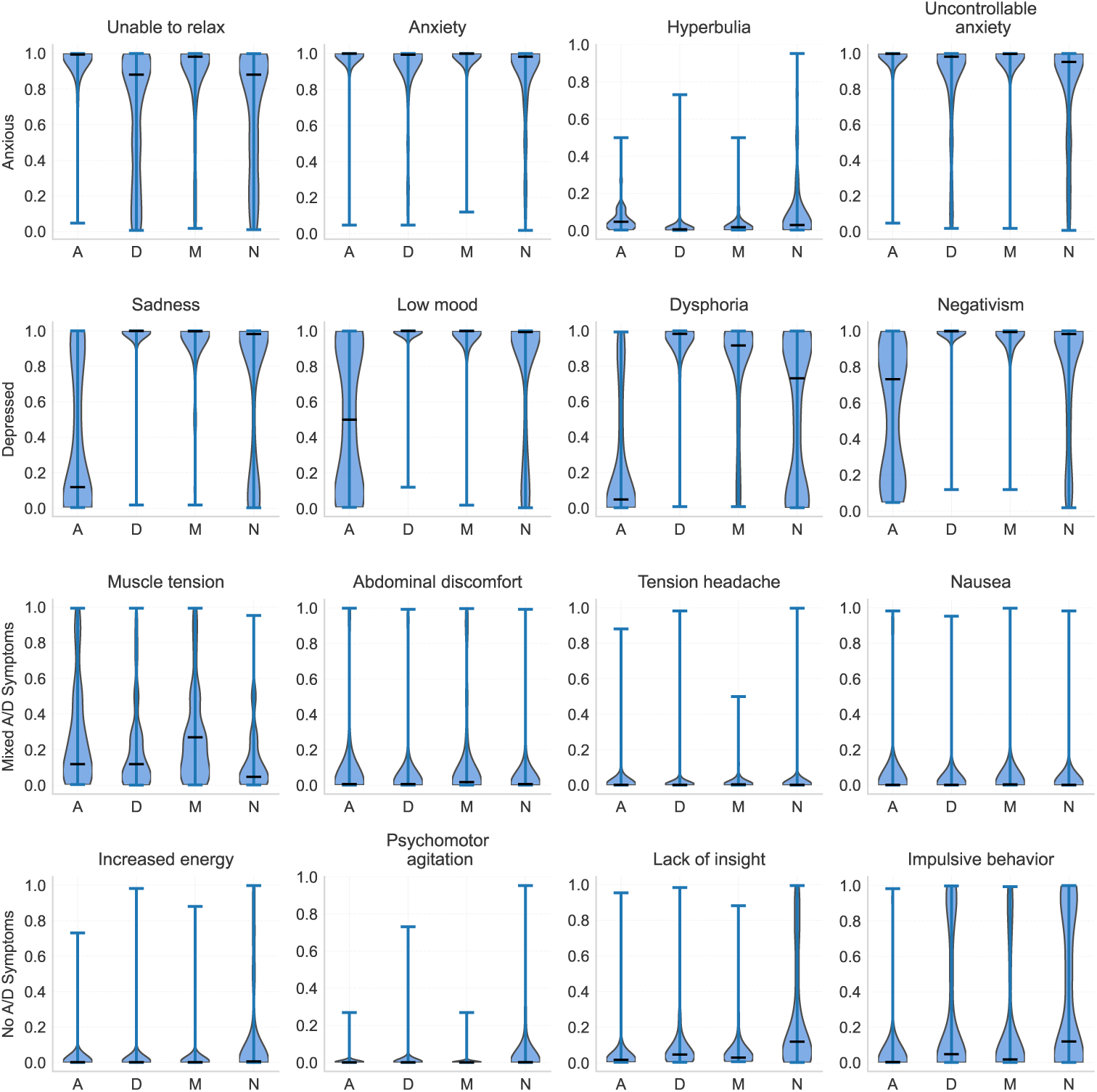
Violin plots of the distribution of top four silent clinical-related features (p-value *<* 0.01) across diagnostic groups (Anxious [A], Depressed [D], Mixed Anxiety/Depression [M], and No Anxiety/Depression [N]). Each feature, identified in the plot titles, was selected based on statistical significance (lowest p-values) within its respective diagnostic group and exhibited higher median values than other groups.

**Fig. A4:**
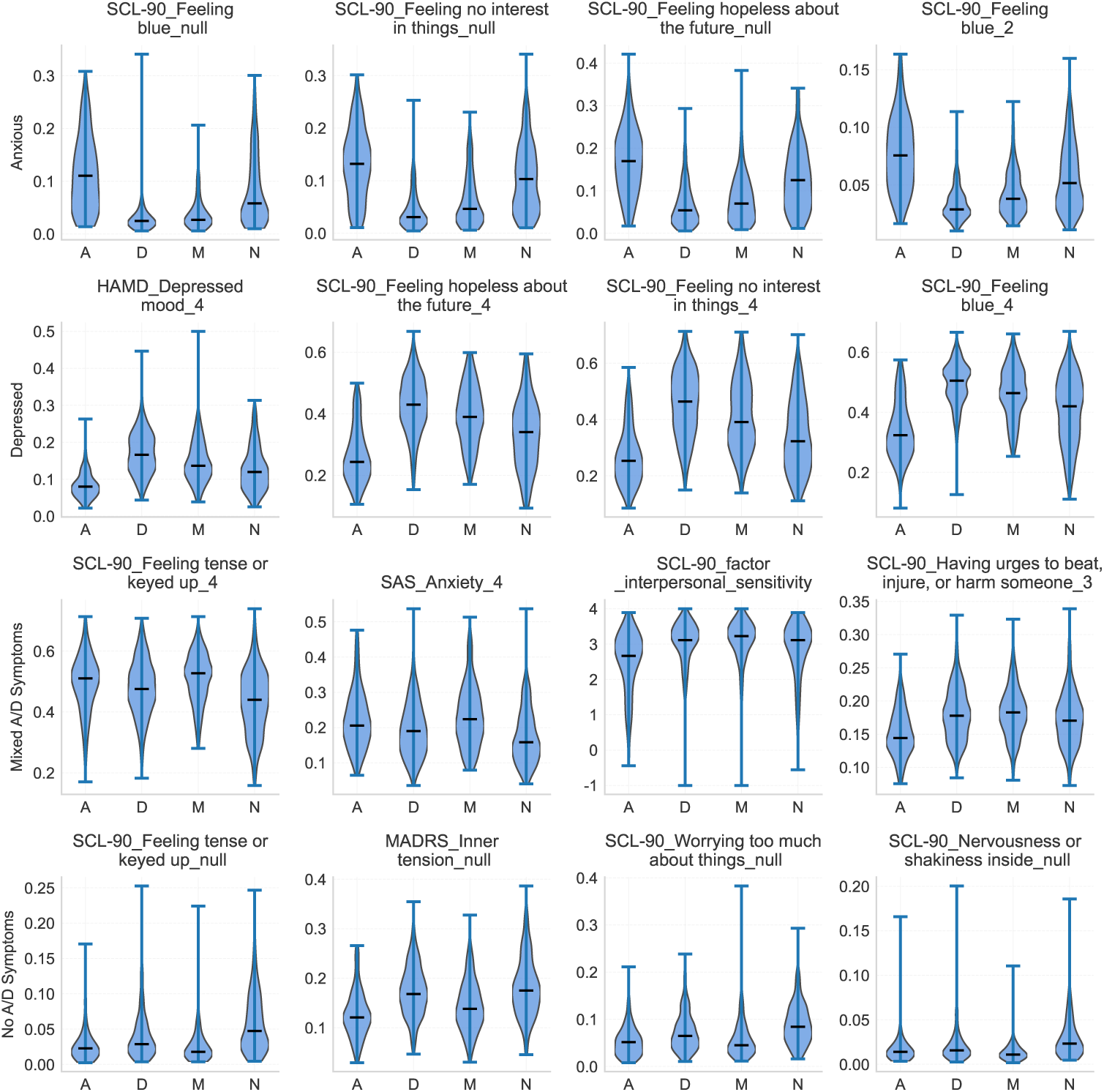
Violin plots of the distribution of four silent assessment-related features (p-value *<* 0.01) across diagnostic groups (Anxious [A], Depressed [D], Mixed Anxiety/Depression [M], and No Anxiety/Depression [N]). Each feature, identified in the plot titles, was selected based on statistical significance (lowest p-values) within its respective diagnostic group and exhibited higher median values than other groups. Feature nomenclature follows the format: Scale_SymptomName_Rating, where a ‘NULL’ rating indicates the absence of symptom identified by LLM.

## Footnotes

1 https://github.com/hiyouga/LLaMA-Factory

2 https://github.com/vllm-project/vllm

3 https://github.com/fxsjy/jieba

4 https://github.com/asweigart/pyaudacity

5 https://github.com/slhck/ffmpeg-normalize

